# Impact of Discoordinated Care on Healthcare Utilization and Survival in Stage IV Non-Small Cell Lung Cancer Patients

**DOI:** 10.1101/2020.12.28.20248904

**Authors:** Samuel P Heilbroner, Eric P Xanthopoulos, Mark A Hoffman, Donna Buono, Ben Y Durkee, Michael Corradetti, Tony JC Wang, Jason D Wright, Alfred I Neugut, Dawn L Hershman, Nicholas C Rohs, Simon K Cheng

## Abstract

**Background:** Cancer patients’ treatment is often dispersed across multiple physician communities that may not exchange information. We measured the association between this type of discoordinated care and healthcare utilization, cost, and mortality in stage IV non-small cell lung cancer (NSCLC).

**Methods:** Stage IV NSCLC patients ≥65 years were identified from the Surveillance, Epidemiology, and End Results database attached to Medicare claims. A discoordinated care subgroup was selected using a novel index that approximated how many physician communities each patient encountered. Communities were defined by (1) using claims data to create a nationwide social network of healthcare providers and then (2) dividing that network into communities using cluster label propagation. Associations between discoordinated care and overall survival (OS), cancer-specific survival (CSS), hospitalizations, the burden of diagnostic imaging, and cost were assessed.

**Results:** Of the 11,417 patients in our cohort, 5,855 received discoordinated care. Discoordination was associated with younger age, higher socioeconomic status, higher physician density, and lack of a home health aide. Discoordinated care was associated with improved OS and CSS (HR = 0.92, 95% CI 0.88 - 0.95 for OS). However, discoordinated patients also received 32% more MRIs (p = 0.007) and paid $494.02 more for imaging (p = 0.004). There was no association with other kinds of imaging, rates of hospitalization, or other healthcare costs, including total cost.

**Conclusions:** Discoordinated care was associated with additional MRIs, but also improved survival. The reason is unclear, but discoordinated patients may be seeking the best care at the expense of continuity.

**Key Points:** *Question:* How does dispersing healthcare across multiple physician communities impact healthcare utilization and survival in patients with advanced stage non-small cell lung cancer?

*Findings:* In this retrospective cohort study, we found that discoordinated care was associated with increased utilization of MRIs and total cost of imaging. Surprisingly, it was also associated with improved survival.

*Meaning:* Cancer patients with dispersed healthcare may be seeking care through a tertiary care center or clinical trial. This may lead to increased healthcare utilization but also improved survival.

## Introduction

Continuity of care is defined as consistent, “seamless” treatment over time involving various healthcare providers and settings.^1^ It has been associated with patient satisfaction,^2^ health promotion,^3^ increased medication adherence,^4^ reduced length of hospital stay,^5^ and improved mortality.^6^ Conversely, discoordinated care is associated with repeated laboratory tests and imaging.^7,8^

Because medical records are difficult to transfer from one physician network to another,^9^ treatment across too many physician communities can represent one type of discoordinated care. But while there is much research on continuity of care on the individual physician level, less is known about continuity at the physician community level.^10^ This type of discoordination is particularly relevant to cancer patients, whose care is complex and often dispersed across multiple oncology centers. Indeed, in other complex conditions like diabetes and heart failure, patients with a closely connected physician team had significantly lower costs of care and fewer hospitalizations.^11^ But the notion that discoordinated care worsens outcomes has not been formally tested in cancer patients. Because of the severity of cancer illness, patients who seek out the best care at the expense of continuity may do better than expected.

To determine the impact of discoordinated care in oncology, we compared the outcomes of a large cohort of elderly stage IV non-small cell lung cancer (NSCLC) patients who received discoordinated care to a similar cohort that received continuous care. Care was categorized as continuous or discoordinated using a novel index that measures how closely members of a patient’s healthcare team are connected. Outcomes included overall survival, cancer-specific survival, burden of diagnostic imaging, number of hospitalizations, and cost.

## Methods

A detailed description of our methods is provided in the appendix and makes use of several references.^5,12-37^ A brief description is provided below:

### Cohort Selection

We studied patients ≥ 65 years old diagnosed with stage IV NSCLC between 2005 and 2012 in the Surveillance, Epidemiology, and End Results database attached to Medicare claims (SEER-Medicare). Only patients with at least two documented provider encounters after diagnosis were included. Continuity was measured during the first 6 months following diagnosis, so patients who lived less than 6 months were excluded. Several other standard selection criteria were used to ensure data quality and completeness. These criteria are ubiquitous in the SEER-Medicare literature and are enumerated in this study’s consort diagram (appendix Table T1).

### Scoring Patient Continuity of Care

To measure a patient’s level of continuity of care we created a novel index based on repeated contact between patients and cohesive physician communities. Physician communities were defined by creating a large physician social network using Medicare claims data and then dividing it into groups using a well-known community detection algorithm. First we compiled a list of all national carrier (NCH) and outpatient facility (OUTSAF) claims filed between 2005 and 2013. From each of these 119,156,351 claims, we extracted (1) the patient who was billed and (2) the billing physician. This information represented 9,834,329 unique doctor-patient relationships. From these pairs, a physician social network was constructed. The 470,569 nodes of the network represented individual physicians and the 118,186,702 connections of the network represented shared patients. Connections were weighted by the number of shared patients. Physicians were divided into communities using cluster label propagation (CLP).^38^ Although there are many community detection algorithms, CLP was the only algorithm that could process such a large network. The process is summarized in appendix Table T2.

Using these physician communities and the previously described Continuity of Care Index,^39^ we created the Care Dispersion Index (CDI) to capture whether a patient received care from a single physician community or many. It is calculated using the formula

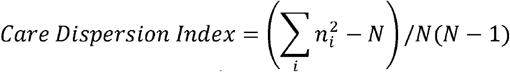

where n_i_ is the number of encounters to the i^th^ physician community and N is the total number of patient encounters. The CDI approximates the probability that two of a patient’s randomly selected encounters are to the same physician-community. Figure 1 gives some examples. Patient encounters were enumerated using NCH and OUTSAF claims from diagnosis to 6 months after diagnosis. Patients with a CDI score above the median were put in the continuous care group, while those below the median were put in the discoordinated care group. We also conducted 6 sensitivity analyses where we varied the CDI cutoff between coordinated and discoordinated care. **Appendix 2** summarizes the results, which generally agreed with what is reported below.

**Figure 1:**
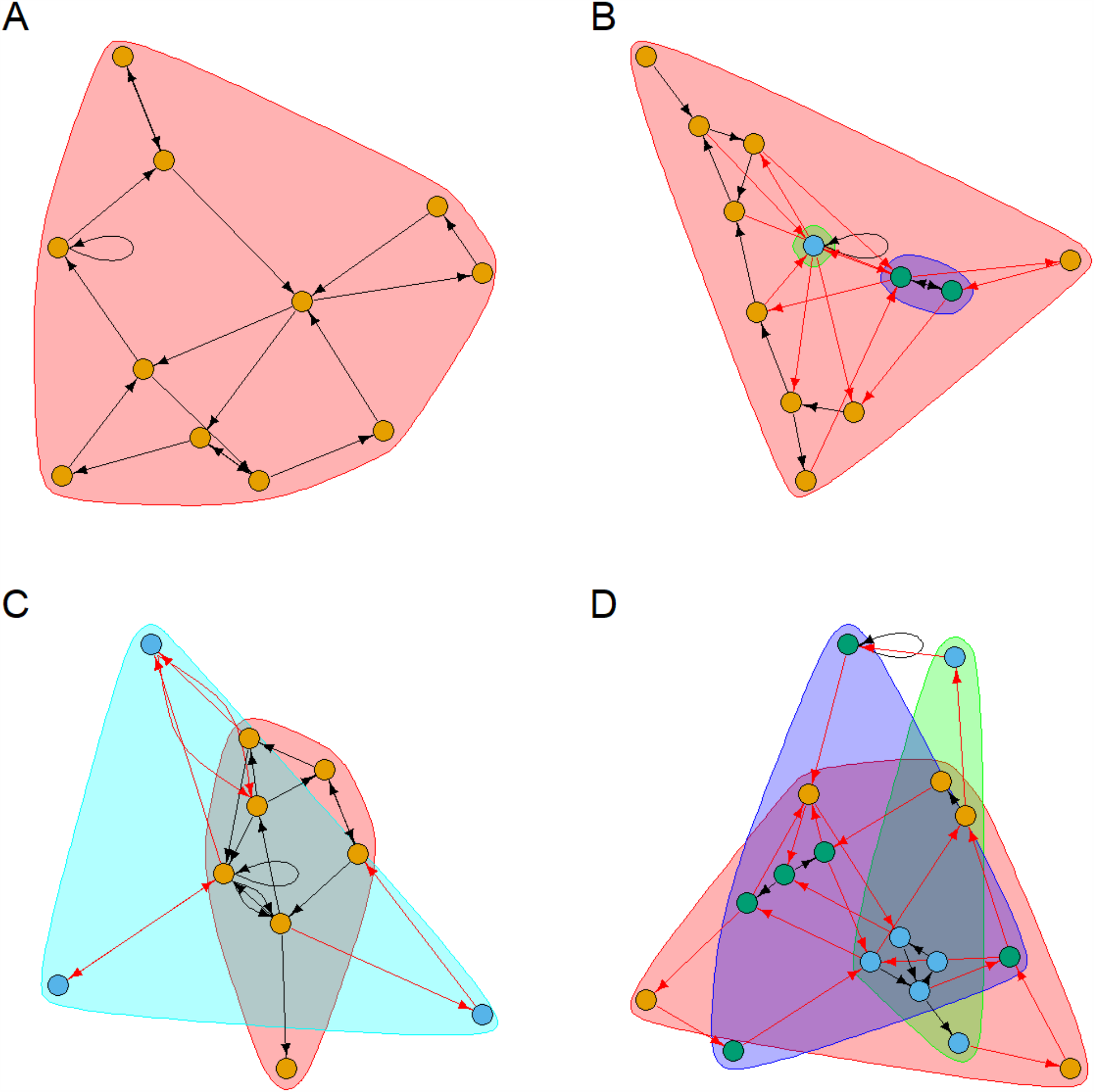
This figure provides some intuition for the CDI. It shows the journey of four simulated patients through the healthcare system. Simulated data was used to adhere with National Cancer Institute guidelines requiring that no real individual patient or physician information be shared in this manuscript. Red arrows indicate when a patient has crossed from one physician community into another. Nodes represent physicians and directed edges represent the sequence of patient visits. Nodes of the same color are from the same physician community. Some patients received all of their care from within a single physician community while others received it from many communities. This notion of continuity versus discoordination is quantified by the CDI. The CDI for the patients listed are A = 1, B =0.65, C = 0.51, D = 0.32. The maximum CDI is 1. As a patient encounters more physician communities, their CDI approaches 0. Abbreviations: CDI, Care Dispersion Index

### Calculating Outcomes

We compared overall survival, cancer-specific survival, the burden of diagnostic imaging, the number of hospitalizations, and the cost of care between the continuous care and discoordinated care subgroups. Overall and cancer-specific survival are available in the SEER-Medicare database. The number of PET, MRI, CT, bone, nuclear medicine, radiographic, and ultrasonic scans were calculated by counting the number of unique HCPC codes recorded in the NCH and OUTSAF files for each type of imaging during (supplementary Table T3).^40^ For each patient, hospitalizations were calculated by counting the number of Medpar records. Adopting the method pioneered by Smith et al,^28^ we calculated the cost of imaging and hospitalizations as well as the total cost of care by summing the total amount reimbursed by Medicare for these services. We also calculated the cost of chemotherapy and radiotherapy treatment using HCPC and ICD-9 codes (appendix section M1-M2 and Table T3).

To help establish a causal relationship between continuity of care and outcomes,^41^ we deliberately measured outcomes during a time period (6-12 months after diagnosis) after continuity was measure (0-6 month after diagnosis). Therefore, this analysis could not assess healthcare utilization and cost during the period immediately following diagnosis. As an exploratory analysis, we also measured the burden of diagnostic imaging and hospitalizations during the initial 6 months after diagnosis. However, because the time period over which continuity and outcomes are measured overlap in the exploratory analysis, these results are susceptible to reverse causality and establish a relationship that is more likely to be associative and not casual (i.e. higher healthcare utilization could cause more care discoordination). In the exploratory analysis, we attempted to account for reverse-causality by matching patients on their total number of patient encounters, but acknowledge that this may not completely account for time-dependent effects.

### Other Covariates

In addition to continuity of care, we included several other covariates in our analyses to account for confounding: age at diagnosis, sex, race, marital status, socioeconomic status stratified by quintile, regional physician density (e.g. physician availability), state, the Charlson comorbidity index,^42^ medical history of COPD, use of supplemental oxygen, billing for home-health aid services, use of different staging procedures, histology, and residence in a rural or urban area. A detailed description of these covariates and how they were calculated is in the appendix section M1. We also included the total number of encounters in the 6 months following diagnosis as a covariate to account for the possibility that sicker patients with more encounters needed to see more specialists, and therefore suffered more discoordinated care.

### Statistical Analysis

Statistical methods are described in detail in the appendix section M3. Covariates associated with discoordinated care were calculated using a logistic model. Using this model, patients were propensity-score matched in a 1:1 fashion without replacement based on receipt of discoordinated care. A bivariate distribution table confirmed that all covariates were well balanced after matching. Overall and cancer-specific survival were assessed using multivariate Cox regressions. Kaplan-Meier plots were generated for the matched and unmatched cohorts. The burden of imaging and hospitalizations were compared by plotting the mean cumulative count^43^ over time and by calculating the average number of events in each group from 6 to 12 months after diagnosis. Finally, for each cost outcome (i.e., the cost of chemotherapy, MRIs etc…), continuous and discoordinated cohorts were compared using the bootstrap method.

## Results

There were 11,417 patients in our cohort. Patients were divided along the median CDI of 0.92 (appendix figure F1). By this definition, 5,562 (48.7%) received continuous care and 5,855 (51.3%) received discoordinated care. We successfully matched 4,548 continuous patients 1:1 with discoordinated counterparts for a total matched cohort size of 9,096. All covariates were well balanced after matching (appendix Table T4). The median follow-up time was 57.6 months in the unmatched cohort and did not vary significantly between the continuous and discoordinated subgroups or between the matched and unmatched cohorts (appendix Table T5).

For each patient, the CDI score was calculated based on his interaction with 1,266 physician communities defined by CLP (modularity = 0.65). The median community was composed of 22 providers but varied widely between 2 and 25,101 providers. The distribution of group sizes was right skewed, with a mean group size of 372.

On logistic regression, discoordinated care was associated with decreasing age (OR 0.79, 95% CI 0.88 - 1.03 for patients 85+ versus 65-75 yrs.), high socioeconomic status (OR 1.34, 95% CI 1.16 - 1.55 for patients in the 5^th^ quintile for SES versus the 1^st^ quintile), physician density (OR = 1.22, 95% CI 1.08 - 1.38 for 4^th^ quartile density versus 1^st^ quartile), residence in several states (Californians and Michiganders had the highest propensity for discoordinated care), use of home health aide services (OR = 1.50, 95% CI 1.08 - 2.09), receipt of a staging fine needle aspiration (OR = 0.92, 95% CI 0.86 - 1.00), and care in a rural area (OR = 0.46, 95% CI = 0.41 - 0.52 for care in an SEER urban area versus a rural area) (Table 1). Discoordination was not well associated with the total number of provider encounters on our logistic model, and was actually inversely correlated in our bivariate distribution analysis (appendix Table T4).

**Table 1:** Logistic model describing factors associated with discoordinated care. Abbreviations: HHA, Home Health Aide; PET, Positron Emission Tomography; FNA, Fine Needle Aspiration; SCC, Squamous Cell Carcinoma; NSCLC NOS, Non-small Cell Lung Cancer Not Otherwise Specified

| **Variable** | **OR (95% CI, p-value)** |
| --- | --- |
| **Age** |  |
| 65 - 75 | Ref |
| 75 - 85 | 0.95 (0.88 - 1.03, 0.242) |
| 85+ | 0.79 (0.68 - 0.93, <0.01) |
| **Sex** |  |
| Male | Ref |
| Female | 1.07 (0.98 - 1.16, 0.114) |
| **Race** |  |
| White | Ref |
| Black | 1.50 (0.74 - 3.05, 0.259) |
| Hispanic | 1.04 (0.55 - 1.95, 0.908) |
| Other | 1.00 (0.69 - 1.46, 0.992) |
| Unknown | 1.26 (0.91 - 1.74, 0.162) |
| **Marital Status** |  |
| Not Married | Ref |
| Married | 1.05 (0.97 - 1.14, 0.208) |
| **Socioeconomic Status** |  |
| 1st quintile | Ref |
| 2nd quintile | 1.13 (0.99 - 1.28, 0.076) |
| 3rd quintile | 1.16 (1.00 - 1.34, 0.044) |
| 4th quintile | 1.15 (1.00 - 1.31, 0.046) |
| 5th quintile | 1.34 (1.16 - 1.55, <0.01) |
| **Physician Density** |  |
| 1st quartile | Ref |
| 2nd quartile | 0.89 (0.74 - 1.08, 0.235) |
| 3rd quartile | 0.63 (0.52 - 0.75, <0.01) |
| 4th quartile | 1.22 (1.08 - 1.38, <0.01) |
| Unknown | 0.84 (0.76 - 0.93, <0.01) |
| **State** |  |
| California | Ref |
| Connecticut | 0.83 (0.69 - 1.00, 0.049) |
| Georgia | 0.78 (0.68 - 0.90, <0.01) |
| Hawaii | 0.30 (0.18 - 0.50, <0.01) |
| Iowa | 0.44 (0.36 - 0.53, <0.01) |
| Kentucky | 0.28 (0.24 - 0.34, <0.01) |
| Louisiana | 0.57 (0.47 - 0.70, <0.01) |
| Michigan | 1.48 (1.23 - 1.77, <0.01) |
| New Jersey | 0.75 (0.66 - 0.86, <0.01) |

Interestingly, discoordinated care was associated with improved overall (HR = 0.92, 95% CI 0.88 - 0.95) and cancer-specific (HR = 0.91, 95% CI 0.87 - 0.95) survival on multivariate Cox regression analysis. Several other covariates were also associated with overall-survival, including advanced age (HR = 1.21, 95% CI 1.11 - 1.31), female sex (HR = 0.85, 95% CI 0.82 - 0.89), Hispanic race (HR = 1.51, 95% CI 1.11 - 2.03), marriage (HR = 0.95, 95% CI 0.91 - 0.99), high socioeconomic status (e.g. HR = 0.85, 95% CI 0.79 - 0.92 for patients in the 5^th^ quintile of SES), a high Charlson comorbidity index (e.g. HR = 1.11, 95% CI 1.03 - 1.20 for an index of 3+), O2 dependence (HR = 1.09, 95% CI 1.04 - 1.14) and a large number of encounters with providers (e.g. HR = 1.13, 95% CI 1.06 - 1.21 for patients in the 4^th^ quartile of number of provider encounters). Residence in Iowa was also associated with poorer outcomes (e.g. HR = 1.19, 95% CI 1.08 - 1.31 for Iowans vs. Californians). Finally, staging procedures (i.e. PET scan, mediastinoscopy, and FNA) were associated with improved survival (e.g. HR = 0.86, 95% CI 0.82 - 0.90 for PET scans) and adenocarcinoma had a better prognosis compared to all other histologies (e.g. HR = 1.09, 95% CI 1.04 - 1.15 for squamous cell carcinoma vs. adenocarcinoma). Results were similar for CSS. Full OS and CSS results are reported in the appendix tables T6-T7.

Patients with discoordinated care also had better OS and CSS on Kaplan-Meier analysis both before and after matching (Figure 2). By study design, all patients in this study lived at least 6 months after diagnosis. After that time, median survivals were 7.6 vs. 8.6 months for matched patients with discoordinated versus continuous care (p < 0.001). Results for CSS and unmatched patients were similar (appendix Table T5).

**Figure 2:**
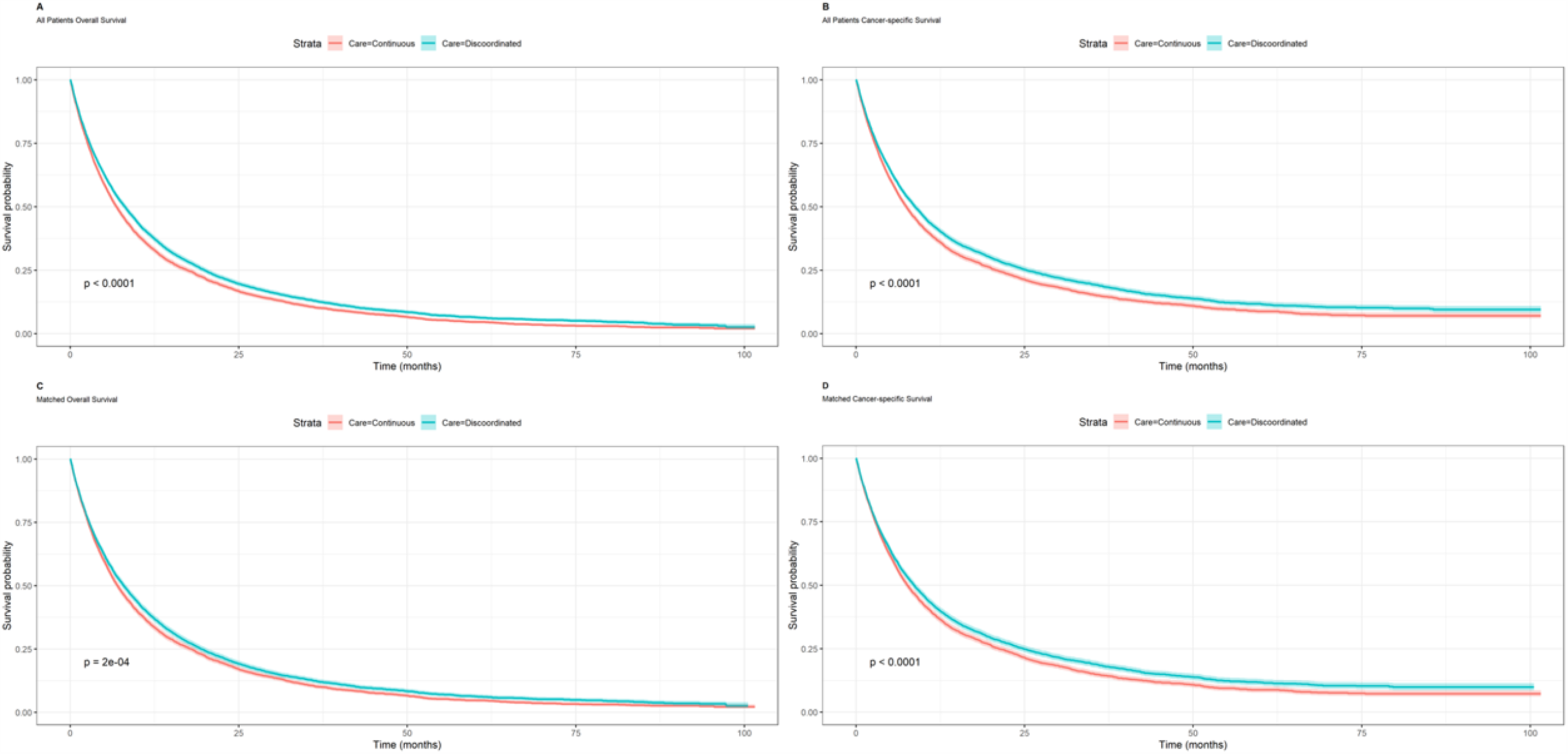
Kaplan-Meier analysis comparing continuous and discoordinated care for (A) overall survival in all patients, (B) cancer-specific survival in all patients, (C) overall survival in matched patients, and (D) cancer-specific survival in matched patients.

Although discoordinated care was associated with better survival, it was also associated with a higher burden of MRIs and overall cost of diagnostic imaging. Discoordinated patients from the matched cohort underwent an additional 0.24 MRI scans (p = 0.007) (Figure 3), a 32% increase. This translated directly into an increase in cost: the average cost of diagnostic imaging was $2,296.43 in continuous patients versus $2,790.45 in discoordinated patients (diff = $494.02, p = 0.004). The number of PET, MRI, CT, bone, nuclear medicine, radiographic, and ultrasonic scans were not significantly associated with continuity of care, although many trended towards increased utilization with discoordinated care (appendix Table T8). The rate of hospitalizations was not increased in discoordinated patients from the matched cohort (p = 0.185). Finally, radiation, chemotherapy, hospitalization associated, and total costs were not associated with continuity (Table 2). Results were generally similar for unmatched patients and are reported in the tables.

**Table 2:**
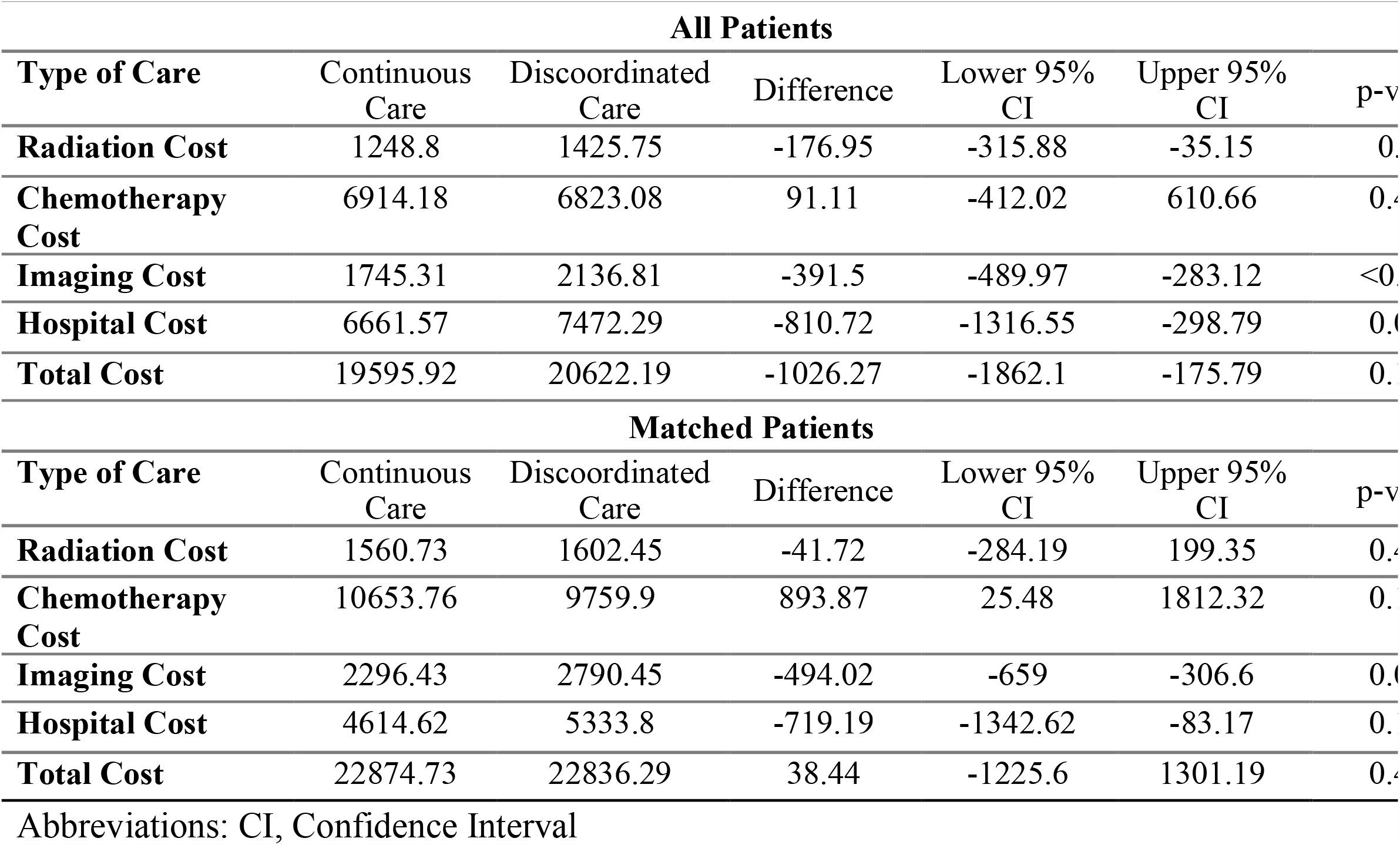
The cost of radiation associated, chemotherapy associated, imaging associated, hospital associated, and overall care. Results are divided by receipt of discoordinated care. Abbreviations: CI, Confidence Interval

**Figure 3:**
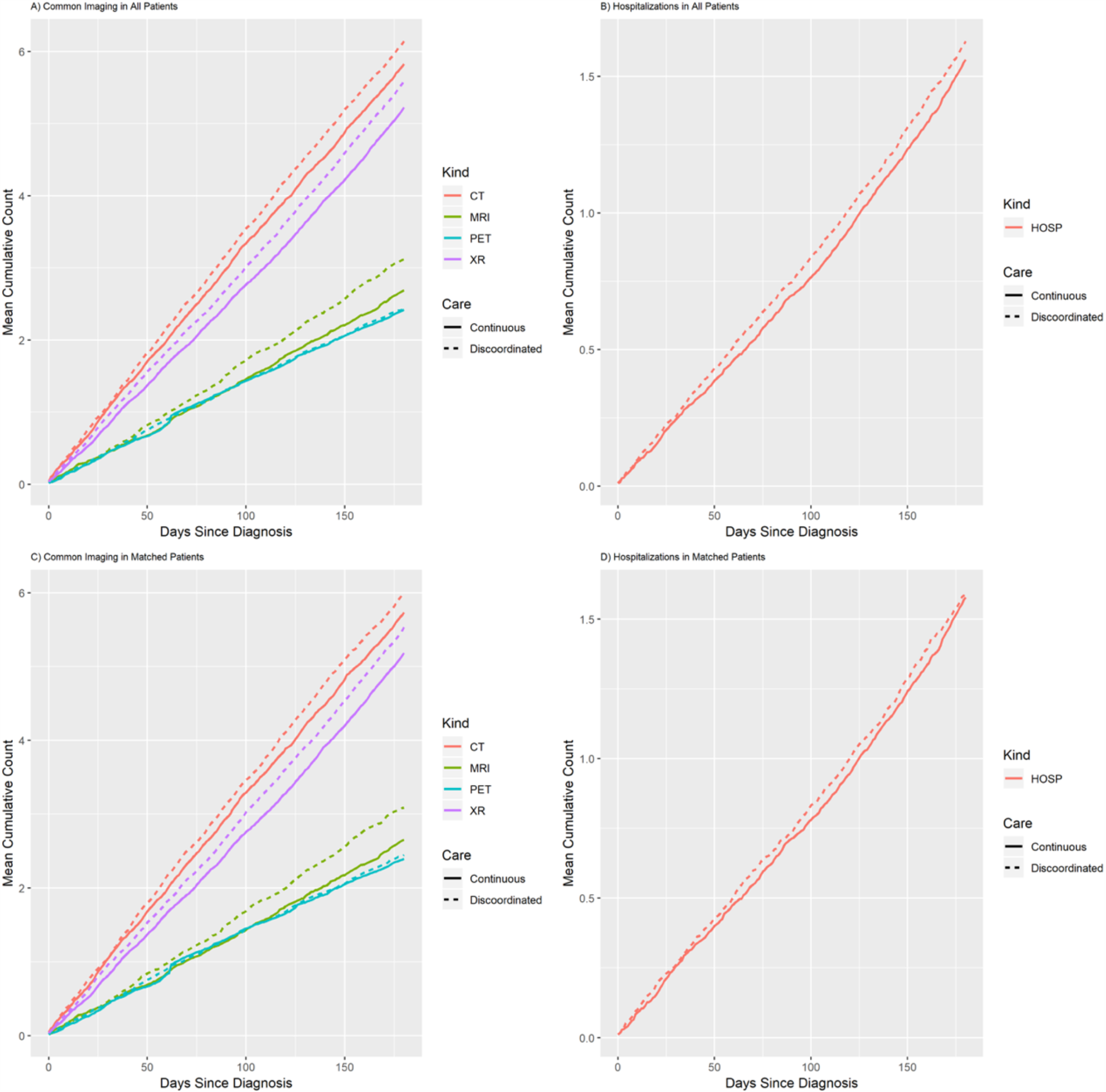
The mean cumulative count of common imaging modalities (A, C) and hospitalizations (B, D) for all (A, B) and matched (C, D) patients. Counts are stratified by whether the patient received continuous or discoordinated care. Cumulative MRIs were significantly associated with the receipt of discoordinated care. Cumulative radiographs, CT scans, and hospitalizations trended towards increased use with discoordinated care, but these results did not achieve significance.

In our time insensitive exploratory analysis, discoordinated care was associated with an increase in almost every type of imaging as well as in the burden of hospitalizations during the first 6 months after diagnosis: 0.42 CT scans (p = 0.034), 0.13 nuclear medicine scans (p = 0.024), 1.24 radiographs (p < 0.001), 0.14 ultrasounds (p = 0.023), and 0.15 hospitalizations (p = 0.006). Discoordinated care was still not associated with the burden of PET scans or bone scans (appendix Table T9).

In the matched cohort of this exploratory analysis, discoordinated care was associated with an additional $803.81 spent on imaging (p < 0.001) and $2963.08 spent on hospitalizations (p < 0.001). Interestingly, discoordinated care was also associated with a reduction in the cost of chemotherapy (Diff = $1,593.41, p = 0.002). Discoordinated care did not affect radiation costs. There was a trend toward increased total cost but this did not achieve significance (Diff = $1600.95, p = 0.1). Results in unmatched patient were similar (appendix Table T10).

## Discussion

We found that discoordinated care was associated with an increase in median overall survival of 1 month in stage IV NSCLC patients. On the other hand, the existing literature suggests that discoordinated care increases mortality.^44^ But of the 22 studies reviewed linking continuity to mortality, none looked at cancer patients. Because of the acuity of cancer illness, discontinuity in this setting may be driven by different factors like a patient’s desire to receive the best care through a tertiary care center or clinical trial. In oncology, the patient who seeks the best care at the expense of continuity may be rewarded by better outcomes. In this way, our finding that discontinuous care improves survival in NSCLC may be analogous to the literature linking increased travel distance to improved survival.^45,46^

There are several other possible explanations for an association between discontinuity and improved survival. Discontinuous care may introduce new insights into the process of diagnosis and management by making patients’ healthcare teams more diverse.^47^ It also seems likely that the ability to go from one facility to another indicates better resources and performance status, which are independently associated with improved survival. Indeed, we found that patients who received discoordinated care were more likely to be young, wealthy, have better access to healthcare (i.e. patients living in an area of high physician density), and function without the need of a home health aide. Patients residing in rural areas were also more likely to receive discoordinated healthcare. Because of the paucity of large integrated physician networks in rural America, these patients may have to seek care across multiple smaller, disjointed, networks.

Although discoordinated care was associated with improved mortality, it was also significantly associated with a 32% increase in MRI scans and a 22% increase in the cost of imaging. In our primary analysis, we did not find a significant association between discoordinated care and an increased burden of other forms of imaging besides MRI or hospitalizations. We also did not find a significant association between continuity and radiation, chemotherapy, hospitalization associated, and total costs (Table 2).

Previous literature has found stronger associations between continuity and healthcare utilization. 3 out of 4 recently reviewed papers found a statistically significant association between continuity and hospitalizations.^48^ Another paper showed reduced hospitalizations with increased continuity in breast, colorectal, and prostate cancer survivors, and used very similar methods to our own.^10^ This discrepancy may represent true differences between stage IV NSCLC patients and survivors of these other cancers. Another possibility is that the power of our study was weakened by measuring imaging and hospitalizations over a distinct time period after we measured continuity. In an exploratory analysis, we showed that the association between continuity and the burden of almost all imaging types and hospitalizations became significant if these outcomes and continuity are measured during the same period. However, these results should be interpreted with caution. Patients with increased healthcare utilization can *de facto* have decreased continuity (e.g. patients are treated by new doctors when they go to the emergency room) or end up being treated by new doctors as a result of resource utilization (e.g. hospitalized care is often provided by several subspecialists).^48^ Such bias is often termed reverse causality or time dependent bias. Although we attempted to control for total healthcare utilization by matching patients on their total number of provider encounters, this may not be sufficient to account for all time-varying effects.

Previous literature linking social network analysis to outcomes has used “care density” to measure continuity of care instead of the CDI.^10,11^ Care density directly measures how often a patient’s doctors share patients. But physicians from the same network do not always share patients; a geriatrician and pediatrician from the same hospital may not, for example. The CDI has the advantage of designating care from within the same physician community as continuous, regardless of whether individual physicians share patients in a pairwise fashion. The CDI has another advantage over care density: it has an obvious application to policy. The physician communities described in this paper could be published and policy makers could provide incentives for patients to receive all of their care from within a single community.

In summary, we found that discontinuous care was associated with improved survival but also increased healthcare utilization. In the context of the previous literature, this study confirms continuities’ impact on utilization while calling into question its effect on mortality, particularly in active cancer patients. The greater complexity of cancer care may explain these discrepancies. Multiple provisions of the Affordable Care Act seek to improve care coordination.^11^ But curbing patient choice may have unintended consequences. Further research is needed to determine whether the potential benefits of continuous care outweigh the costs.

## Data Availability

SEER-Medicare data is available through the NCI website.

