## Appendix 1 for "Impact of Discoordinated Care on Healthcare Utilization and Survival in Stage IV Non-Small Cell Lung Cancer Patients"

| **Page #** | **Supplementary Section Number** | **Title** |
| --- | --- | --- |
| 2 | M1 | Description and calculation of covariates. |
| 2 | M2 | Calculating cost of care |
| 2-3 | M3 | Statistical analysis |
| 4 | T1 | Consort Diagram for Cohort in Main Study |
| 5 | T2 | Consort diagram describing construction of physician communities |
| 6-7 | T3 | Billing codes used to calculate covariates and outcomes |
| 8-9 | T4 | Bivariate distribution of patient characteristics |
| 10 | T5 | Median overall survival, cancer-specific survival, and follow-up time for matched and unmatched patients. |
| 11-12 | T6 | Cox regression output for overall survival |
| 13-14 | T7 | Cox regression output for cancer-specific survival |
| 15 | T8 | Mean number of images/hospitalizations in main analysis from 6 to 12 months after diagnosis. |
| 16 | T9 | Mean number of images/hospitalizations in exploratory analysis from 0 to 6 months after diagnosis. |
| 17 | T10 | Mean cost of care in exploratory analysis from 0 to 6 months after diagnosis. |
| 18 | F1 | Distribution of Team Based Continuity of Care Indices |
| 19 | F2 | Plots of the mean cumulative count for imaging and hospitalizations in exploratory analysis |
| 20 | R1 | References for the Appendix |

**Supplementary Section M1**.	Description and calculation of covariates

For each patient, several additional covariates were collected or calculated. We collected patient demographic and tumor information, including age at diagnosis, sex, race, marital status, and primary tumor histology from SEER registry data. Whether a patient resided in a rural or urban area was also collected.

We calculated an aggregate socioeconomic status score with education, poverty level, and income information from the 2010 census tract data, as described previously by Du et al.^1^ Using the method pioneered by Smith et. al, we calculated total physician density by geographic area.^2^ The physician density represents the ratio of the number of doctors in a given area to the total population, and is a proxy for physician availability.

Using Medicare claims in the 12 months prior to cancer diagnosis, we calculated the Klabunde adaptation of the Charlson comorbidity index to assess the prevalence of comorbid disease in our cohort.^3-6^ We separately used the Centers for Medicaid & Medicare Services (CMS) Chronic Conditions Data Warehouse (CCW) algorithms to identify chronic obstructive pulmonary disease (COPD).^7^ Use of supplemental oxygen was flagged in patients with a DME bill for oxygen in the 6 months prior to diagnosis (**Supplementary Table T3**). Using Medicare MEDPAR, OUTSAF, NCH, HHA, and DME claims codes from 1.5 months before diagnosis to 1 month after diagnosis (**Supplementary Table T3**), we tabulated the use of staging procedures (e.g. PET scans, Mediastinoscopies, and Fine Needle Aspirations). In a similar fashion, we recorded receipt of radiation and chemotherapy in the first 6 months after diagnosis (**Supplementary Table T3**).^8^^,^^9^

**Supplementary Section M2**. Calculating cost of care

Total costs included Medicare payment aggregated from inpatient facility claims in the Part A Medicare Provider Analysis and Review (MEDPAR) files, outpatient facility claims in the Part B hospital-based Outpatient Claims (OUTSAF) files, and the in- or outpatient physician claims in the Part B Carrier Claims (formerly the Physician/Supplier or NCH) files. Narrower radiotherapy and chemotherapy costs summed claims based on the codes previously described (**Supplemental Table T3**).

We adjusted costs for inflation, normalizing them to the year 2019 using the Prospective Pricing Index for Part A claims and the Medicare Economic Index for Part B claims. We simultaneously adjusted costs for geographic variation using the geographic adjustment factor for Part A claims and the Geographic Practice Cost index for Part B claims. The National Cancer Institute’s Health Services and Economics Branch of the Applied Research Program provided all the adjustments used to tabulate costs.

**Supplementary Section M3**. Statistical analysis

*Chi-square bivariate analysis***.** We compared the distribution of patient characteristics between the two treatment groups with the Pearson’s chi-square test. All cell sizes in this table were > 5, obviating the need for Fischer’s exact test. To protect patient anonymity and consistent with policies governing the use of SEER-Medicare data, we did not report subgroups with a size less than 11.

*Logistic regression testing associations with Discoordinated Care***.** We included all covariates in a multivariable logistic regression model to predict discoordinated care. We assessed the quality of our model by checking the area under the curve (c = 0.64) and the Hosmer Lemeshow goodness of fit (p = 0.35 > 0.05).

*Propensity score matching***.** Based on the results of our logistic regression, we created a propensity score model to account for confounding by our covariates. Patients were randomly sorted and then matched 1-to-1 without replacement to a nearest neighbor with a match caliper of 0.05. Bivariate association p-values showed that all covariates were well balanced after matching.

*Kaplan-Meier analysis testing association between Discoordinated Care and Survival***.** The association between discoordinated care and survival were assessed using the Kaplan-Meier method with censorship at the earliest of the following: death, or the end of the study period on December 31, 2013. For each endpoint, the proportional hazards assumption with respect to discoordinated care was tested visually by inspection of log-log plots and analytically using Schoenfield residuals. Plots were generated for matched and unmatched cohorts.

*Cox regression testing associations with Survival***.** As in our logistic model, Cox regressions were created using all covariates and receipt of discoordinated care as inputs. For all survival endpoints, the proportional hazards assumption with respect to discoordinated care was satisfied, and goodness- of-fit for all final models was acceptable (p > 0.05).

*Estimating the Difference in Cost and Healthcare Utilization*. A non-parametric bootstrap model used 1,000 samples to estimate the 95% confidence interval (CI) around the mean cost differences between the continuous and discoordinated care groups. We estimated these differences for both the original, unmatched cohort and the 1-to-1 matched sub-cohort. This same technique was used to detect differences in the number of images or hospitalizations between the continuous and discoordinated care groups.

The burden of healthcare utilization was described visually for common imaging modalities and hospitalizations using plots of the mean cumulative count over time.^10^

*Statistical Software*

Analyses were performed with SAS version 9.4 and R version 3.5.0.^11^ Analyses in R made use of the following packages: ff,^12^ data.table,^13^ microbenchmark,^14^ lubridate,^15^ bit64,^16^ ggplot2,^17^ survival,^18^^,^^19^ survminer,^20^ labelled,^21^ tableone,^22^ MatchIt,^23^ dtplyr,^24^ magrittr,^25^ Publish,^26^ stringr,^27^ boot,^28^ dplyr,^29^ ggrepel,^30^ pROC,^31^ ResourceSelection,^32^ haven,^33^ and igraph.^34^

| Supplementary Table T1. Consort Diagram for Cohort in Main Study | |
| --- | --- |
| Selection Criteria | **# of Patients** |
| All Lung Cancer Patients in the SEER-Medicare Database | 600,828 |
| Only Select SEER-Records for First Cancer Diagnosis | 548,939 |
| 1st Cancer is of the Lung | 471,916 |
| Reporting Source should not be autopsy or death certificate. | 459,716 |
| Age of diagnosis should be greater than 65. | 381,444 |
| Original or current reason for entitlement should be age. | 379,047 |
| Delete if date of death between SEER and Medicare is off by > 3 months. | 378,158 |
| Take only cases diagnosed from 2005 to 2012 | 165,421 |
| Exclude members of an HMO 12 months before to 12 months after diagnosis. | 120,780 |
| Have both Medicare Part A & B Coverage 12 months before to 12 months after diagnosis. | 106,377 |
| Non-small cell lung cancer | 76,873 |
| Stage IV | 30,032 |
| Calculatable CDI (at least 2 provider encounters) | 29,481 |
| Survival > 6 months | 11,417 |
| Matched | 9,096 |
| Abbreviations: SEER, Surveillance, Epidemiology, and End Results Database; HMO, Health Maintenance Organization; CDI, Care Dispersion Index | |

| **Supplementary Table T2.** Consort diagram describing construction of physician communities. | |
| --- | --- |
| **Selection Criteria** | **Result** |
| All PHYSICIAN Carrier (NCH) and Outpatient Facility (OUTSAF) Claims from 2005 to 2013 from the SEER-Medicare Database | 119,156,351 claims |
| Exclude claims with a recorded UPIN and NPI that are not from the same provider according to the SEER-Medicare Crosswalk file | 109,789,618 claims |
| Delete claims without a recorded UPIN or NPI. Use Crosswalk file to create one physician ID from UPIN and NPI. | 108,930,575 claims |
| Keep one record for each unique physician-patient interaction. | 9,834,329 doctor-patient relationships |
| Social network of providers who share patients. Nodes represent physicians. Edges are weighted by the number of shared patients between providers. | A social network of 470,569 doctors connected by 118,186,702 physician-physician relationships |
| Group physicians in social network into communities using weighted cluster label propagation. | Social network as above but now divided into 1,266 communities |
| Abbreviations: UPIN, Unique Physician Identification Number; NPI, National Provider Identifier | |

| **Supplementary Table T3.** Billing codes for used to calculate covariates and outcomes in this analysis. | | | | |
| --- | --- | --- | --- | --- |
| **Treatment** | **ICD-9 Codes** | **CPT/HCPCS Codes** | **Revenue Center/**  **Diagnosis Related Codes** | **DME File MTUSIND Code** |
| Chemotherapy | 99.25,  V58.1, V66.2, V67.2  E9331, E9307 | Q0083, Q0084, Q0085, 96400-96599, J9000-J9999, G0355-G0362 | 0331, 0332, 0335  410 |  |
| Radiation | V58.0, V66.1, V67.1  92.21, 92.22, 92.23, 92.24, 92.25, 92.26, 92.27, 92.28, 92.29 | 77401-77499, 77520-77525, 77750-77799 | 0330, 0333 |  |
| Supplemental O2 |  |  |  | 4 |
| Diagnostic PET scan |  | 78810-78816, G0030-G0047, G0210-G0235, G0125-G0126, G0163-G0165, G0252-G0254, G0296, G0330, G0331, G0336 |  |  |
| Diagnostic Mediastinoscopy | 3422 | 39400 |  |  |
| Diagnostic FNA | 3326-3327 | 32405, 10021-10022 |  |  |
| CT |  | 70460, 71250, 71260, 71270, 72192-72194, 74150, 74160, 74170, 76380, 70496, 70498, 71275, 72191, 73206, 73706, 74175, 75635, 70450-70492, 73200- 73202, 73700-73702, 70450-70492 |  |  |
| MRI |  | 71550-71552, 72196-72197, 74182-74183, 76094, 71555, 72198, 73725, 75552-75564, 70336, 70540-70559, 73219-73223, 73719-73723 |  |  |
| PET |  | 78810-78816, G0126, G0164-G0165, G211-G228, G232-G234, G214-G215, G227-G228, G0253-G0254, G0330-G0331, 78491-78492, G31-G47, 78608-78609, G0336 |  |  |
| Radiographs |  | 74000-74022, 74291, 74415, 76000-76001, 76125, 71011-71035, 74220, 74241-74280, 76091-76092, 76150, G203-G207, 70031-70130, 70140, 70160, 70191-70260, 70310, 70328, 70360, 70371, 71100-71130, 72011-72120, 72190, 72202, 73000, 73020, 73050, 73070, 73090, 73110, 73130, 73500, 73520, 73550, 73562, 73565, 73600, 73620, 73650, 76006, 76040, 76062 |  |  |
| Nuclear Medicine Scan |  | 78428-78483, 78496, 78001-78075, 78103-78104, 78195, 78202-78264, 78290, 78301-78320, 78580-78601, 78606-78607, 78615, 78635, 78647, 78660, 78701-78724, 78727-78761 |  |  |
| Bone Scan |  | 76070-76071, 76075-77083, 78350-78351, G0130-G0133 |  |  |
| Ultrasound |  | 76506, 76511-76604, 76700, 76770, 76776, 76800, 76831, 76857, 76872-76873, 76970, 93875, 93881-93888, 93891-93893, 93923-93931, 93970-93990 |  |  |
| Abbreviations: ICD-9, International Classification of Diseases, 9th Revision, Clinical Modification (ICD-9-CM); CPT/HCPCS, Current Procedural Terminology/ Healthcare Common Procedure Coding System; CT, Computer Tomography; MRI, Magnetic Resonance Imaging; PET, Positron Emission Tomography | | | | |

| Supplementary Table T4. Bivariate distribution of patient characteristics across all patients and matched patients. | | | | | | |
| --- | --- | --- | --- | --- | --- | --- |
|  | All Patients | | | Matched Patients | | |
|  | **Continuous** | **Discoordinated** | **p-value** | **Continuous** | **Discoordinated** | **p-value** |
| n | 5562 | 5855 |  | 4548 | 4548 |  |
| Age (%) |  |  | 0.123 |  |  | 0.759 |
| 65 - 75 | 2994 (53.8) | 3191 (54.5) |  | 2437 (53.6) | 2448 (53.8) |  |
| 75 - 85 | 2161 (38.9) | 2292 (39.1) |  | 1787 (39.3) | 1794 (39.4) |  |
| 85+ | 407 (7.3) | 372 (6.4) |  | 324 (7.1) | 306 (6.7) |  |
| Sex = Female (%) | 2765 (49.7) | 2995 (51.2) | 0.129 | 2270 (49.9) | 2309 (50.8) | 0.426 |
| Race (%) |  |  | <0.001 |  |  | 0.607 |
| White | > 4700 (85) | > 4884 (83) |  | > 3831 (84) | > 3801 (84) |  |
| Black | 513 (9.2) | 489 (8.4) |  | 419 (9.2) | 414 (9.1) |  |
| Hispanic | 41 (0.7) | 72 (1.2) |  | 41 (0.9) | 42 (0.9) |  |
| Other | 296 (5.3) | 398 (6.8) |  | 245 (5.4) | 279 (6.1) |  |
| Unknown | <12 (0.2) | <12 (0.2) |  | <12 (0.2) | <12 (0.2) |  |
| Marital Status = Married (%) | 2959 (53.2) | 3208 (54.8) | 0.092 | 2454 (54.0) | 2458 (54.0) | 0.95 |
| Socioeconomic Status (%) |  |  | <0.001 |  |  | 0.988 |
| 1st quintile | 959 (17.2) | 854 (14.6) |  | 704 (15.5) | 695 (15.3) |  |
| 2nd quintile | 1140 (20.5) | 1138 (19.4) |  | 921 (20.3) | 930 (20.4) |  |
| 3rd quintile | 877 (15.8) | 934 (16.0) |  | 712 (15.7) | 720 (15.8) |  |
| 4th quintile | 1440 (25.9) | 1482 (25.3) |  | 1183 (26.0) | 1165 (25.6) |  |
| 5th quintile | 1146 (20.6) | 1447 (24.7) |  | 1028 (22.6) | 1038 (22.8) |  |
| Physician Density (%) |  |  | <0.001 |  |  | 0.318 |
| 1st quartile | 2265 (40.7) | 1823 (31.1) |  | 1599 (35.2) | 1582 (34.8) |  |
| 2nd quartile | 818 (14.7) | 1272 (21.7) |  | 767 (16.9) | 841 (18.5) |  |
| 3rd quartile | 1068 (19.2) | 1373 (23.5) |  | 1022 (22.5) | 989 (21.7) |  |
| 4th quartile | 1182 (21.3) | 1267 (21.6) |  | 1048 (23.0) | 1017 (22.4) |  |
| Unknown | 229 (4.1) | 120 (2.0) |  | 112 (2.5) | 119 (2.6) |  |
| State (%) |  |  | <0.001 |  |  | 0.881 |
| California | 1374 (24.7) | 2019 (34.5) |  | 1358 (29.9) | 1362 (29.9) |  |
| Connecticut | 267 (4.8) | 395 (6.7) |  | 261 (5.7) | 273 (6.0) |  |
| Georgia | 696 (12.5) | 688 (11.8) |  | 569 (12.5) | 583 (12.8) |  |
| Hawaii | 85 (1.5) | 37 (0.6) |  | 26 (0.6) | 37 (0.8) |  |
| Iowa | 361 (6.5) | 266 (4.5) |  | 215 (4.7) | 238 (5.2) |  |
| Kentucky | 711 (12.8) | 291 (5.0) |  | 316 (6.9) | 291 (6.4) |  |
| Louisiana | 367 (6.6) | 295 (5.0) |  | 287 (6.3) | 269 (5.9) |  |
| Michigan | 396 (7.1) | 530 (9.1) |  | 396 (8.7) | 380 (8.4) |  |
| New Jersey | 807 (14.5) | 826 (14.1) |  | 706 (15.5) | 689 (15.1) |  |
| New Mexico | 84 (1.5) | 124 (2.1) |  | 82 (1.8) | 85 (1.9) |  |
| Utah | 66 (1.2) | 56 (1.0) |  | 52 (1.1) | 52 (1.1) |  |
| Washington | 348 (6.3) | 328 (5.6) |  | 280 (6.2) | 289 (6.4) |  |
| Charlson Score (%) |  |  | 0.82 |  |  | 0.677 |
| 0 | 3383 (60.8) | 3563 (60.9) |  | 2766 (60.8) | 2792 (61.4) |  |
| 1-2 | 1773 (31.9) | 1848 (31.6) |  | 1452 (31.9) | 1415 (31.1) |  |
| 3+ | 406 (7.3) | 444 (7.6) |  | 330 (7.3) | 341 (7.5) |  |
| COPD = Yes (%) | 2185 (39.3) | 2178 (37.2) | 0.023 | 1734 (38.1) | 1738 (38.2) | 0.948 |
| O2 dependent = Yes (%) | 1286 (23.1) | 1292 (22.1) | 0.185 | 1017 (22.4) | 1025 (22.5) | 0.86 |
| HHA = Yes (%) | 65 (1.2) | 96 (1.6) | 0.04 | 58 (1.3) | 57 (1.3) | 1 |
| Diag. PET Scan = Yes (%) | 3748 (67.4) | 3845 (65.7) | 0.055 | 3033 (66.7) | 3028 (66.6) | 0.929 |
| Diag. Mediastinoscopy = Yes (%) | 176 (3.2) | 214 (3.7) | 0.164 | 154 (3.4) | 158 (3.5) | 0.863 |
| Diag. FNA = Yes (%) | 2634 (47.4) | 2702 (46.1) | 0.202 | 2145 (47.2) | 2138 (47.0) | 0.9 |
| Histology (%) |  |  | 0.001 |  |  | 0.994 |
| Adenocarcinoma | 2714 (48.8) | 3092 (52.8) |  | 2302 (50.6) | 2292 (50.4) |  |
| SCC | 1241 (22.3) | 1200 (20.5) |  | 980 (21.5) | 988 (21.7) |  |
| Large Cell | 188 (3.4) | 169 (2.9) |  | 145 (3.2) | 143 (3.1) |  |
| Other | 232 (4.2) | 219 (3.7) |  | 180 (4.0) | 174 (3.8) |  |
| NSCLC, NOS | 1187 (21.3) | 1175 (20.1) |  | 941 (20.7) | 951 (20.9) |  |
| Rural vs. Urban = Urban (%) | 4676 (84.1) | 4789 (81.8) | 0.001 | 3808 (83.7) | 3763 (82.7) | 0.217 |
| # of Provider Encounters (%) |  |  | 0.001 |  |  | 0.624 |
| 1st quartile | 694 (12.5) | 737 (12.6) |  | 583 (12.8) | 563 (12.4) |  |
| 2nd quartile | 1055 (19.0) | 1162 (19.8) |  | 905 (19.9) | 903 (19.9) |  |
| 3rd quartile | 1534 (27.6) | 1762 (30.1) |  | 1279 (28.1) | 1333 (29.3) |  |
| 4th quartile | 2279 (41.0) | 2194 (37.5) |  | 1781 (39.2) | 1749 (38.5) |  |
| Abbreviations: HHA, Home Health Aide; PET, Positron Emission Tomography; FNA, Fine Needle Aspiration; SCC, Squamous Cell Carcinoma; NSCLC NOS, Non-small Cell Lung Cancer Not Otherwise Specified | | | | | | |

| Supplementary Table T5. Median overall survival, cancer-specific survival, and follow-up time for matched and unmatched patients. | | | |
| --- | --- | --- | --- |
| All Patients | | | |
| Statistic | Continuous | Discoordinated | All |
| Overall Survival | 6.85 | 8.07 | 7.45 |
| Cancer-specific Survival | 7.45 | 8.73 | 8.07 |
| Follow-up Time | 56.56 | 57.55 | 57.55 |
| Matched Cohort | | | |
| Statistic | Continuous | Discoordinated | All |
| Overall Survival | 6.99 | 7.91 | 7.45 |
| Cancer-specific Survival | 7.61 | 8.63 | 8.07 |
| Follow-up Time | 56.56 | 58.57 | 57.55 |

| Supplementary Table T6. Cox regression output for overall survival | | |
| --- | --- | --- |
| Variable | **Univariate HR (95% CI, p-value)** | **Multivariate HR (95% CI, p-value)** |
| Continuity of Care |  |  |
| Continuous | Ref | Ref |
| Discoordinated | 0.90 (0.86 - 0.93, <0.01) | 0.92 (0.88 - 0.95, <0.01) |
| Age |  |  |
| 65 - 75 | Ref | Ref |
| 75 - 85 | 1.05 (1.01 - 1.09, 0.02) | 1.06 (1.02 - 1.11, <0.01) |
| 85+ | 1.18 (1.09 - 1.28, <0.01) | 1.21 (1.11 - 1.31, <0.01) |
| Sex |  |  |
| Male | Ref | Ref |
| Female | 0.84 (0.81 - 0.87, <0.01) | 0.85 (0.82 - 0.89, <0.01) |
| Race |  |  |
| White | Ref | Ref |
| Black | 1.25 (0.90 - 1.75, 0.185) | 1.38 (0.99 - 1.93, 0.058) |
| Hispanic | 1.36 (1.01 - 1.84, 0.043) | 1.51 (1.11 - 2.03, <0.01) |
| Other | 1.32 (1.11 - 1.58, <0.01) | 1.27 (1.06 - 1.52, <0.01) |
| Unknown | 1.11 (0.94 - 1.30, 0.211) | 1.13 (0.96 - 1.32, 0.142) |
| Marital Status |  |  |
| Not Married | Ref | Ref |
| Married | 0.97 (0.93 - 1.01, 0.099) | 0.95 (0.91 - 0.99, 0.023) |
| Socioeconomic Status |  |  |
| 1st quintile | Ref | Ref |
| 2nd quintile | 0.95 (0.89 - 1.01, 0.12) | 0.95 (0.89 - 1.02, 0.143) |
| 3rd quintile | 0.88 (0.82 - 0.94, <0.01) | 0.89 (0.83 - 0.96, <0.01) |
| 4th quintile | 0.86 (0.81 - 0.91, <0.01) | 0.89 (0.83 - 0.95, <0.01) |
| 5th quintile | 0.82 (0.77 - 0.87, <0.01) | 0.85 (0.79 - 0.92, <0.01) |
| Physician Density |  |  |
| 1st quartile | Ref | Ref |
| 2nd quartile | 1.03 (0.95 - 1.11, 0.498) | 0.99 (0.90 - 1.09, 0.832) |
| 3rd quartile | 1.10 (1.03 - 1.17, <0.01) | 0.95 (0.86 - 1.04, 0.230) |
| 4th quartile | 1.05 (0.99 - 1.10, 0.085) | 1.02 (0.96 - 1.09, 0.436) |
| Unknown | 0.97 (0.93 - 1.02, 0.220) | 0.99 (0.94 - 1.04, 0.600) |
| State |  |  |
| California | Ref | Ref |
| Connecticut | 1.08 (0.99 - 1.18, 0.083) | 1.06 (0.96 - 1.17, 0.224) |
| Georgia | 1.15 (1.08 - 1.23, <0.01) | 1.09 (1.02 - 1.18, 0.016) |
| Hawaii | 1.17 (0.97 - 1.41, 0.102) | 1.28 (1.00 - 1.63, 0.046) |
| Iowa | 1.25 (1.14 - 1.36, <0.01) | 1.19 (1.08 - 1.31, <0.01) |
| Kentucky | 1.17 (1.08 - 1.25, <0.01) | 1.07 (0.98 - 1.17, 0.114) |
| Louisiana | 1.20 (1.10 - 1.31, <0.01) | 1.12 (1.02 - 1.24, 0.020) |
| Michigan | 1.09 (1.01 - 1.17, 0.035) | 1.09 (1.00 - 1.20, 0.060) |
| New Jersey | 1.05 (0.99 - 1.12, 0.128) | 1.04 (0.97 - 1.11, 0.275) |
| New Mexico | 1.10 (0.95 - 1.27, 0.207) | 0.99 (0.85 - 1.16, 0.931) |
| Utah | 1.14 (0.95 - 1.38, 0.167) | 1.10 (0.91 - 1.33, 0.329) |
| Washington | 1.08 (0.99 - 1.17, 0.097) | 1.06 (0.96 - 1.16, 0.244) |
| Charlson Score |  |  |
| 0 | Ref | Ref |
| 1-2 | 1.09 (1.05 - 1.14, <0.01) | 1.05 (1.01 - 1.10, 0.024) |
| 3+ | 1.19 (1.10 - 1.28, <0.01) | 1.11 (1.03 - 1.20, <0.01) |
| COPD |  |  |
| No | Ref | Ref |
| Yes | 1.10 (1.05 - 1.14, <0.01) | 1.02 (0.98 - 1.07, 0.357) |
| O2 dependent |  |  |
| No | Ref | Ref |
| Yes | 1.13 (1.08 - 1.18, <0.01) | 1.09 (1.04 - 1.14, <0.01) |
| HHA |  |  |
| No | Ref | Ref |
| Yes | 1.12 (0.95 - 1.31, 0.17) | 1.03 (0.87 - 1.21, 0.725) |
| Diag. PET Scan |  |  |
| No | Ref | Ref |
| Yes | 0.86 (0.82 - 0.89, <0.01) | 0.86 (0.82 - 0.90, <0.01) |
| Diag. Mediastinoscopy |  |  |
| No | Ref | Ref |
| Yes | 0.85 (0.77 - 0.95, <0.01) | 0.87 (0.78 - 0.96, <0.01) |
| Diag. FNA |  |  |
| No | Ref | Ref |
| Yes | 0.93 (0.90 - 0.97, <0.01) | 0.94 (0.91 - 0.98, <0.01) |
| Histology |  |  |
| Adenocarcinoma | Ref | Ref |
| SCC | 1.17 (1.11 - 1.23, <0.01) | 1.09 (1.04 - 1.15, <0.01) |
| Large Cell | 1.27 (1.14 - 1.42, <0.01) | 1.19 (1.06 - 1.33, <0.01) |
| Other | 1.10 (0.99 - 1.22, 0.065) | 1.08 (0.97 - 1.19, 0.148) |
| NSCLC, NOS | 1.23 (1.17 - 1.29, <0.01) | 1.20 (1.14 - 1.26, <0.01) |
| Rural vs. Urban |  |  |
| Rural | Ref | Ref |
| Urban | 0.91 (0.86 - 0.96, <0.01) | 1.00 (0.94 - 1.06, 0.994) |
| # of Provider Encounters |  |  |
| 1st quartile | Ref | Ref |
| 2nd quartile | 0.94 (0.88 - 1.01, 0.069) | 0.97 (0.91 - 1.05, 0.477) |
| 3rd quartile | 0.95 (0.89 - 1.02, 0.149) | 1.00 (0.94 - 1.07, 0.908) |
| 4th quartile | 1.08 (1.02 - 1.15, 0.015) | 1.13 (1.06 - 1.21, <0.01) |
| Abbreviations: CI, Confidence Interval; HHA, Home Health Aide; PET, Positron Emission Tomography; FNA, Fine Needle Aspiration; SCC, Squamous Cell Carcinoma; NSCLC NOS, Non-small Cell Lung Cancer Not Otherwise Specified | | |

| Supplementary Table T7. Cox regression output for cancer-specific survival | | |
| --- | --- | --- |
| Variable | **Univariate HR (95% CI, p-value)** | **Multivariate HR (95% CI, p-value)** |
| Continuity of Care |  |  |
| Continuous | Ref | Ref |
| Discoordinated | 0.89 (0.85 - 0.92, <0.01) | 0.91 (0.87 - 0.95, <0.01) |
| Age |  |  |
| 65 - 75 | Ref | Ref |
| 75 - 85 | 1.03 (0.99 - 1.07, 0.193) | 1.05 (1.00 - 1.09, 0.040) |
| 85+ | 1.11 (1.02 - 1.21, 0.012) | 1.14 (1.05 - 1.25, <0.01) |
| Sex |  |  |
| Male | Ref | Ref |
| Female | 0.84 (0.80 - 0.87, <0.01) | 0.85 (0.82 - 0.89, <0.01) |
| Race |  |  |
| White | Ref | Ref |
| Black | 1.22 (0.85 - 1.74, 0.29) | 1.36 (0.94 - 1.95, 0.099) |
| Hispanic | 1.38 (1.00 - 1.91, 0.05) | 1.53 (1.11 - 2.12, 0.010) |
| Other | 1.29 (1.06 - 1.56, 0.01) | 1.25 (1.03 - 1.52, 0.024) |
| Unknown | 1.09 (0.92 - 1.30, 0.32) | 1.12 (0.94 - 1.33, 0.212) |
| Marital Status |  |  |
| Not Married | Ref | Ref |
| Married | 0.99 (0.95 - 1.03, 0.56) | 0.96 (0.92 - 1.01, 0.102) |
| Socioeconomic Status |  |  |
| 1st quintile | Ref | Ref |
| 2nd quintile | 0.97 (0.90 - 1.03, 0.31) | 0.96 (0.89 - 1.03, 0.250) |
| 3rd quintile | 0.89 (0.83 - 0.96, <0.01) | 0.90 (0.83 - 0.97, <0.01) |
| 4th quintile | 0.88 (0.82 - 0.94, <0.01) | 0.90 (0.84 - 0.97, <0.01) |
| 5th quintile | 0.84 (0.78 - 0.90, <0.01) | 0.87 (0.80 - 0.94, <0.01) |
| Physician Density |  |  |
| 1st quartile | Ref | Ref |
| 2nd quartile | 1.03 (0.95 - 1.12, 0.461) | 0.99 (0.90 - 1.09, 0.855) |
| 3rd quartile | 1.08 (1.01 - 1.16, 0.032) | 0.92 (0.84 - 1.01, 0.094) |
| 4th quartile | 1.06 (1.00 - 1.12, 0.038) | 1.04 (0.97 - 1.11, 0.267) |
| Unknown | 0.98 (0.94 - 1.03, 0.411) | 1.00 (0.95 - 1.06, 0.887) |
| State |  |  |
| California | Ref | Ref |
| Connecticut | 1.09 (0.99 - 1.19, 0.079) | 1.07 (0.97 - 1.19, 0.164) |
| Georgia | 1.14 (1.06 - 1.22, <0.01) | 1.09 (1.01 - 1.18, 0.026) |
| Hawaii | 1.17 (0.96 - 1.43, 0.121) | 1.30 (1.00 - 1.68, 0.049) |
| Iowa | 1.27 (1.16 - 1.39, <0.01) | 1.22 (1.10 - 1.35, <0.01) |
| Kentucky | 1.18 (1.09 - 1.28, <0.01) | 1.10 (1.00 - 1.21, 0.044) |
| Louisiana | 1.23 (1.12 - 1.35, <0.01) | 1.17 (1.05 - 1.29, <0.01) |
| Michigan | 1.07 (0.99 - 1.16, 0.107) | 1.12 (1.01 - 1.23, 0.033) |
| New Jersey | 1.06 (0.99 - 1.13, 0.107) | 1.06 (0.98 - 1.13, 0.138) |
| New Mexico | 1.12 (0.96 - 1.30, 0.164) | 1.02 (0.87 - 1.20, 0.815) |
| Utah | 1.19 (0.97 - 1.45, 0.087) | 1.13 (0.93 - 1.39, 0.225) |
| Washington | 1.08 (0.99 - 1.19, 0.090) | 1.07 (0.97 - 1.19, 0.172) |
| Charlson Score |  |  |
| 0 | Ref | Ref |
| 1-2 | 1.05 (1.01 - 1.10, 0.022) | 1.02 (0.98 - 1.07, 0.369) |
| 3+ | 1.07 (0.98 - 1.16, 0.118) | 1.02 (0.94 - 1.11, 0.583) |
| COPD |  |  |
| No | Ref | Ref |
| Yes | 1.06 (1.01 - 1.10, 0.01) | 1.00 (0.95 - 1.04, 0.849) |
| O2 dependent |  |  |
| No | Ref | Ref |
| Yes | 1.09 (1.04 - 1.14, <0.01) | 1.07 (1.02 - 1.13, <0.01) |
| HHA |  |  |
| No | Ref | Ref |
| Yes | 1.11 (0.93 - 1.32, 0.23) | 1.07 (0.89 - 1.27, 0.472) |
| Diag. PET Scan |  |  |
| No | Ref | Ref |
| Yes | 0.84 (0.81 - 0.88, <0.01) | 0.85 (0.81 - 0.89, <0.01) |
| Diag. Mediastinoscopy |  |  |
| No | Ref | Ref |
| Yes | 0.83 (0.74 - 0.93, <0.01) | 0.84 (0.75 - 0.95, <0.01) |
| Diag. FNA |  |  |
| No | Ref | Ref |
| Yes | 0.92 (0.88 - 0.96, <0.01) | 0.94 (0.90 - 0.98, <0.01) |
| Histology |  |  |
| Adenocarcinoma | Ref | Ref |
| SCC | 1.17 (1.10 - 1.23, <0.01) | 1.10 (1.04 - 1.16, <0.01) |
| Large Cell | 1.34 (1.19 - 1.50, <0.01) | 1.25 (1.12 - 1.41, <0.01) |
| Other | 1.10 (0.99 - 1.23, 0.084) | 1.07 (0.96 - 1.20, 0.198) |
| NSCLC, NOS | 1.28 (1.22 - 1.35, <0.01) | 1.25 (1.18 - 1.32, <0.01) |
| Rural vs. Urban |  |  |
| Rural | Ref | Ref |
| Urban | 0.90 (0.85 - 0.95, <0.01) | 0.99 (0.93 - 1.05, 0.710) |
| # of Provider Encounters |  |  |
| 1st quartile | Ref | Ref |
| 2nd quartile | 0.90 (0.84 - 0.97, <0.01) | 0.94 (0.88 - 1.02, 0.133) |
| 3rd quartile | 0.91 (0.85 - 0.97, <0.01) | 0.96 (0.89 - 1.03, 0.239) |
| 4th quartile | 1.04 (0.98 - 1.11, 0.23) | 1.09 (1.02 - 1.17, <0.01) |
| Abbreviations: CI, Confidence Interval; HHA, Home Health Aide; PET, Positron Emission Tomography; FNA, Fine Needle Aspiration; SCC, Squamous Cell Carcinoma; NSCLC NOS, Non-small Cell Lung Cancer Not Otherwise Specified | | |

| Supplementary Table T8. Mean number of images/hospitalizations in main analysis from 6 to 12 months after diagnosis. | | | | | | |
| --- | --- | --- | --- | --- | --- | --- |
| All Patients | | | | | | |
| Event | Continuous Care | Discoordinated Care | Difference | Lower 95% CI | Upper 95% CI | p-value |
| PET Scans | 0.87 | 0.87 | -0.01 | -0.12 | 0.08 | 0.452 |
| MRI Scans | 0.75 | 1.03 | -0.28 | -0.36 | -0.2 | <0.001 |
| CT Scans | 4.36 | 4.74 | -0.38 | -0.62 | -0.16 | 0.045 |
| Bone Scans | 0.02 | 0.04 | -0.02 | -0.03 | -0.01 | 0.055 |
| Nuclear Medicine Scans | 0.36 | 0.36 | 0.01 | -0.04 | 0.05 | 0.442 |
| Radiographs | 3.31 | 3.59 | -0.28 | -0.51 | -0.04 | 0.126 |
| Ultrasounds | 0.54 | 0.61 | -0.07 | -0.14 | -0.02 | 0.115 |
| Hospitalizations | 0.46 | 0.5 | -0.04 | -0.09 | 0 | 0.185 |
| Matched Cohort | | | | | | |
| Event | Continuous Care | Discoordinated Care | Difference | Lower 95% CI | Upper 95% CI | p-value |
| PET Scans | 0.91 | 0.88 | 0.03 | -0.11 | 0.13 | 0.399 |
| MRI Scans | 0.76 | 1 | -0.24 | -0.33 | -0.14 | 0.007 |
| CT Scans | 4.3 | 4.58 | -0.28 | -0.54 | -0.02 | 0.14 |
| Bone Scans | 0.02 | 0.04 | -0.02 | -0.03 | 0 | 0.106 |
| Nuclear Medicine Scans | 0.36 | 0.35 | 0.01 | -0.04 | 0.07 | 0.406 |
| Radiographs | 3.22 | 3.6 | -0.38 | -0.64 | -0.13 | 0.066 |
| Ultrasounds | 0.56 | 0.57 | -0.01 | -0.08 | 0.05 | 0.437 |
| Hospitalizations | 0.45 | 0.49 | -0.04 | -0.09 | 0.01 | 0.185 |
| Abbreviations: CI, Confidence Interval; PET, Positron Emission Tomography; MRI, Magnetic Resonance Imaging; CT, Computer Tomography | | | | | | |

| Supplementary Table T9. Mean number of images/hospitalizations in exploratory analysis from 0 to 6 months after diagnosis. Results are susceptible to effects of reverse causality. | | | | | | |
| --- | --- | --- | --- | --- | --- | --- |
| All Patients | | | | | | |
| Event | Continuous Care | Discoordinated Care | Difference | Lower 95% CI | Upper 95% CI | p-value |
| PET Scans | 1.59 | 1.63 | -0.04 | -0.1 | 0.02 | 0.245 |
| MRI Scans | 1.63 | 1.95 | -0.32 | -0.41 | -0.23 | 0.001 |
| CT Scans | 6.78 | 7.09 | -0.31 | -0.52 | -0.1 | 0.072 |
| Bone Scans | 0.03 | 0.03 | -0.01 | -0.02 | 0 | 0.232 |
| Nuclear Medicine Scans | 0.87 | 0.98 | -0.11 | -0.17 | -0.05 | 0.034 |
| Radiographs | 8.29 | 9.3 | -1.01 | -1.27 | -0.72 | <0.001 |
| Ultrasounds | 0.86 | 1.03 | -0.16 | -0.22 | -0.1 | 0.002 |
| Hospitalizations | 1.15 | 1.25 | -0.11 | -0.15 | -0.06 | 0.018 |
| Matched Cohort | | | | | | |
| Event | Continuous Care | Discoordinated Care | Difference | Lower 95% CI | Upper 95% CI | p-value |
| PET Scans | 1.57 | 1.64 | -0.07 | -0.13 | 0 | 0.17 |
| MRI Scans | 1.63 | 1.96 | -0.33 | -0.43 | -0.23 | <0.001 |
| CT Scans | 6.64 | 7.06 | -0.42 | -0.63 | -0.2 | 0.034 |
| Bone Scans | 0.03 | 0.03 | 0 | -0.01 | 0 | 0.341 |
| Nuclear Medicine Scans | 0.86 | 0.99 | -0.13 | -0.2 | -0.07 | 0.024 |
| Radiographs | 8.17 | 9.41 | -1.24 | -1.53 | -0.93 | <0.001 |
| Ultrasounds | 0.88 | 1.02 | -0.14 | -0.21 | -0.07 | 0.023 |
| Hospitalizations | 1.12 | 1.27 | -0.15 | -0.2 | -0.09 | 0.006 |
| Abbreviations: CI, Confidence Interval; PET, Positron Emission Tomography; MRI, Magnetic Resonance Imaging; CT, Computer Tomography | | | | | | |

| Supplementary Table T10. Mean cost of care in exploratory analysis from 0 to 6 months after diagnosis. Results are susceptible to effects of reverse causality. | | | | | | |
| --- | --- | --- | --- | --- | --- | --- |
| All Patients | | | | | | |
| Type of Care | Continuous Care | Discoordinated Care | Difference | Lower 95% CI | Upper 95% CI | p-value |
| Radiation Cost | 3784.02 | 3942.76 | -158.74 | -387.89 | 64.95 | 0.244 |
| Chemotherapy Cost | 12212.32 | 10869.63 | 1342.69 | 777.85 | 1901.05 | 0.01 |
| Imaging Cost | 4461.23 | 5214.6 | -753.37 | -909.59 | -599.16 | <0.001 |
| Hospital Cost | 12994.86 | 16150.37 | -3155.5 | -3904.25 | -2436.06 | <0.001 |
| Total Cost | 39728.6 | 41442.33 | -1713.73 | -2772.1 | -693.34 | 0.047 |
| Matched Patients | | | | | | |
| Type of Care | Continuous Care | Discoordinated Care | Difference | Lower 95% CI | Upper 95% CI | p-value |
| Radiation Cost | 3733.73 | 3980.51 | -246.78 | -507.3 | 5.95 | 0.152 |
| Chemotherapy Cost | 12275.37 | 10681.96 | 1593.41 | 953.31 | 2251.68 | 0.002 |
| Imaging Cost | 4420.08 | 5223.89 | -803.81 | -962.99 | -634.52 | <0.001 |
| Hospital Cost | 13041.58 | 16004.66 | -2963.08 | -3769.69 | -2185.31 | <0.001 |
| Total Cost | 39613.57 | 41214.52 | -1600.95 | -2722.51 | -454.19 | 0.1 |
| Abbreviations: CI, Confidence Interval | | | | | | |

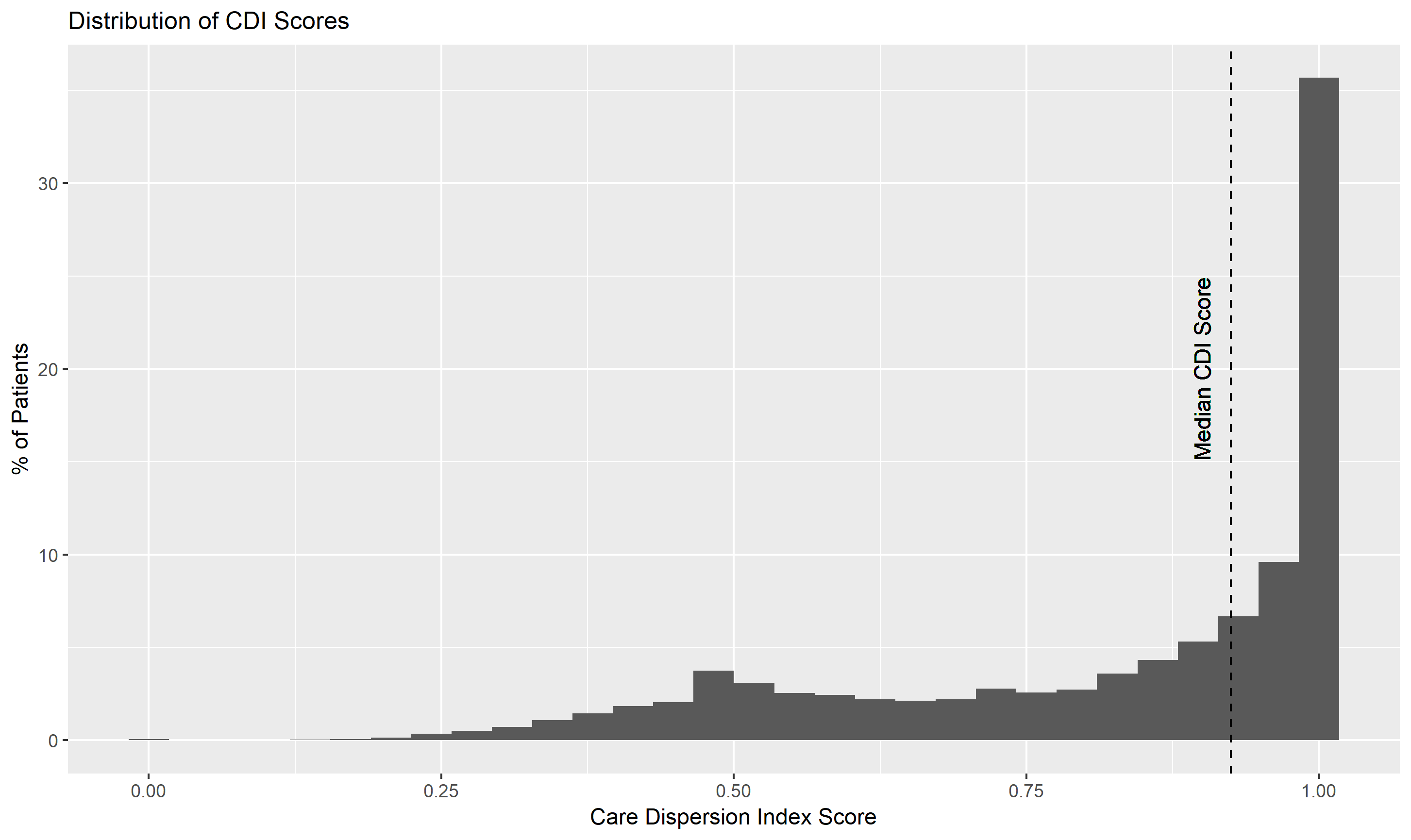

**Supplementary Figure F1.** Distribution of Team Based Continuity of Care Indices (CDI) for Patients in our cohort. Continuous patients had a CDI above the median and discoordinated patient has a CDI below the median.

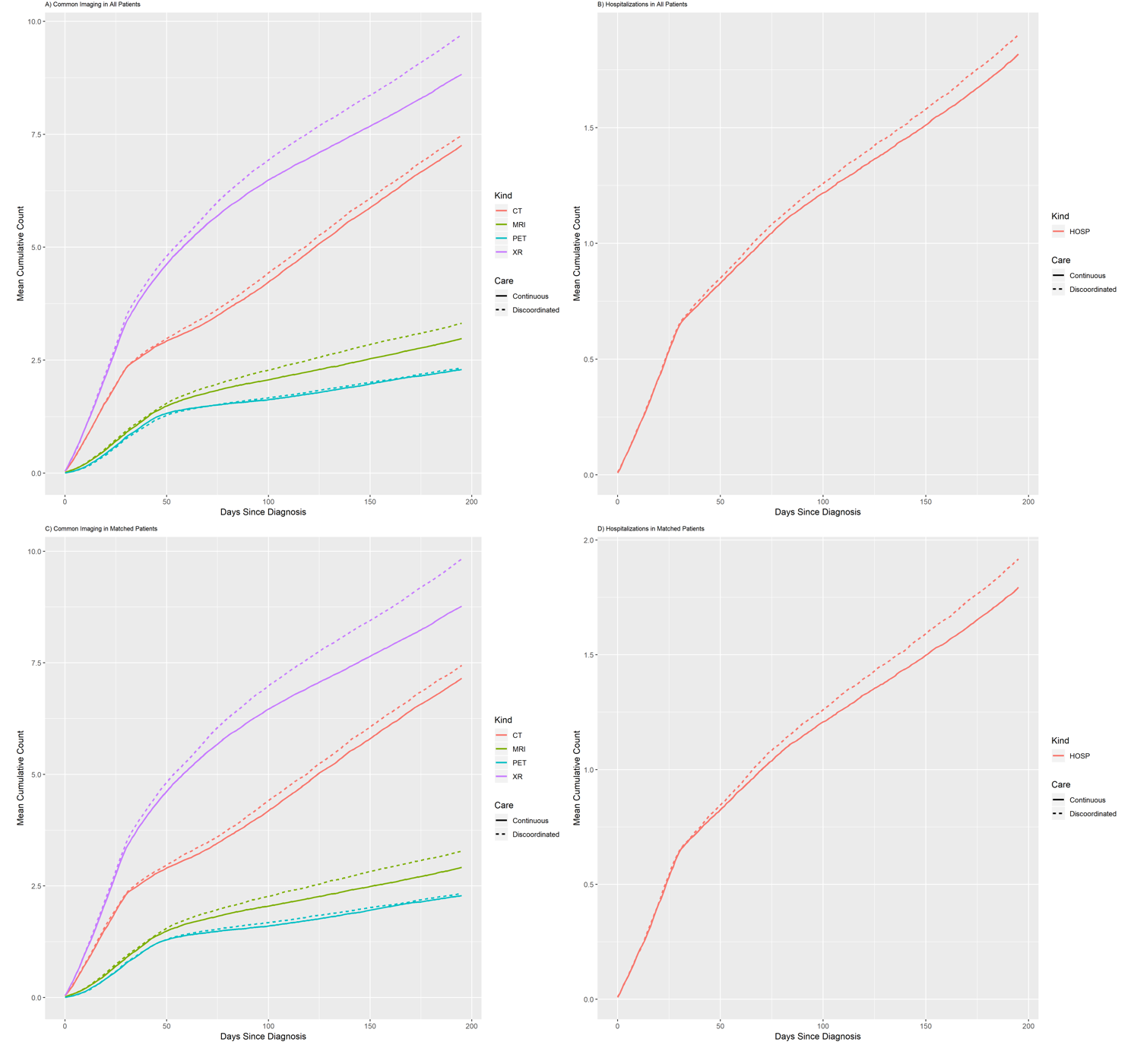

**Supplementary Figure F2**. Plots of the mean cumulative count for imaging and hospitalizations occurring in the first 6 months after Diagnosis. These are the results of our exploratory analysis and are susceptible to reverse causality bias. P-values are reported in supplementary table T9.

**REFRENCES FOR THE APPENDIX**

1.	Du XL, Fang S, Vernon SW, et al. Racial disparities and socioeconomic status in association with survival in a large population‐based cohort of elderly patients with colon cancer. *Cancer: Interdisciplinary International Journal of the American Cancer Society.* 2007;110(3):660-669.

2.	Smith BD, Pan I-W, Shih Y-CT, et al. Adoption of intensity-modulated radiation therapy for breast cancer in the United States. *Journal of the National Cancer Institute.* 2011;103(10):798-809.

3.	<https://healthcaredelivery.cancer.gov/seermedicare/program/comorbidity.html>. NCIDoCCPSS-MCoCWa.

11.	Team RC. R: A Language and Environment for Statistical Computing. 2018.

12.	Adler D, Gl_ser C, Nenadic O, Oehlschl_gel J, Zucchini W. ff: Memory-Efficient Storage of Large Data on Disk and Fast Access Functions. 2018.

13.	Dowle M, Srinivasan A. data.table: Extension of `data.frame`. 2018.

14.	Mersmann O. microbenchmark: Accurate Timing Functions. 2018.

15.	Grolemund G, Wickham H. Dates and Times Made Easy with lubridate. *Journal of Statistical Software.* 2011;40(3):1-25.

16.	Oehlschl_gel J. bit64: A S3 Class for Vectors of 64bit Integers. 2017.

17.	Wickham H. *ggplot2: Elegant Graphics for Data Analysis.* Springer-Verlag New York; 2016.

18.	Therneau TM. A Package for Survival Analysis in S. 2015.

19.	Therneau TM, Grambsch PM. *Modeling Survival Data: Extending the Cox Model.* Springer; 2000.

20.	Kassambara A, Kosinski M. survminer: Drawing Survival Curves using 'ggplot2'. 2018.

21.	Larmarange J. labelled: Manipulating Labelled Data. 2019.

22.	Yoshida K, Bohn J. tableone: Create 'Table 1' to Describe Baseline Characteristics. 2018.

23.	Ho DE, Imai K, King G, Stuart EA. MatchIt: Nonparametric Preprocessing for Parametric Causal Inference. *Journal of Statistical Software.* 2011;42(8):1-28.

24.	Wickham H. dtplyr: Data Table Back-End for 'dplyr'. 2017.

25.	Bache SM, Wickham H. magrittr: A Forward-Pipe Operator for R. 2014.

26.	Gerds TA, Ozenne B. Publish: Format Output of Various Routines in a Suitable Way for Reports and Publication. 2018.

27.	Wickham H. stringr: Simple, Consistent Wrappers for Common String Operations. 2018.

28.	Canty A, Ripley BD. boot: Bootstrap R (S-Plus) Functions. 2017.

29.	Wickham H, Fran_ois R, Henry L, M_ller K. dplyr: A Grammar of Data Manipulation. 2018.

30.	Slowikowski K. ggrepel: Automatically Position Non-Overlapping Text Labels with 'ggplot2'. 2018.

31.	Robin X, Turck N, Hainard A, et al. pROC: an open-source package for R and S+ to analyze and compare ROC curves. *BMC Bioinformatics.* 2011;12:77-77.

32.	Lele SR, Keim JL, Solymos P. ResourceSelection: Resource Selection (Probability) Functions for Use-Availability Data. 2019.

33.	Wickham H, Miller E. haven: Import and Export 'SPSS', 'Stata' and 'SAS' Files. 2018.

34.	Csardi G, Nepusz T. The igraph software package for complex network research. *InterJournal.* 2006;Complex Systems:1695-1695.
